# The impact of positive surgical margin parameters and pathological stage on biochemical recurrence after radical prostatectomy: a systematic review and meta-analysis

**DOI:** 10.1101/2024.03.21.24304691

**Authors:** Hong Guo, Lei Zhang, Yuan Shao, Kunyang An, Caoyang Hu, Xuezhi Liang, Dongwen Wang

## Abstract

**Background:** To systematically review and perform a meta-analysis on the predictive value of the primary Gleason grade (PGG) at the positive surgical margin (PSM), length of PSM, number of PSMs, and pathological stage of the primary tumor on biochemical recurrence (BCR) in patients with prostate cancer (PCa) after radical prostatectomy (RP).

**Methods:** A systematic literature search was performed using electronic databases, including PubMed, EMBASE, Cochrane Library, and Web of Science, from January 1, 2005, to October 1, 2023. The protocol was pre-registered in PROSPERO. Subgroup analyses were performed according to the different treatments and study outcomes. Pooled hazard ratios with 95% confidence intervals were extracted from multivariate analyses, and a fixed or random effect model was used to pool the estimates. Subgroup analyses were performed to explore the reasons for the heterogeneity.

**Results:** Thirty studies that included 46,572 patients with PCa were eligible for this meta-analysis. The results showed that, compared to PGG3, PGG4/5 was associated with a significantly increased risk of BCR. Compared with PSM ≤3 mm, PSM ≥3 mm was associated with a significantly increased risk of BCR. Compared with unifocal PSM, multifocal PSM (mF-PSM) was associated with a significantly increased risk of BCR. In addition, pT >2 was associated with a significantly increased risk of BCR compared to pT2. Notably, the findings were found to be reliable based on the sensitivity and subgroup analyses.

**Conclusions:** PGG at the PSM, length of PSM, number of PSMs, and pathological stage of the primary tumor in patients with PCa were found to be associated with a significantly increased risk of BCR. Thus, patients with these factors should be treated differently in terms of receiving adjunct treatment and more frequent monitoring. Large-scale, well-designed prospective studies with longer follow-up periods are needed to validate the efficacy of these risk factors and their effects on patient responses to adjuvant and salvage therapies and other oncological outcomes.

## Introduction

Prostate cancer (PCa) is the second most common malignancy among men worldwide ^[1]^, and radical prostatectomy (RP) is the most effective treatment for localized PCa ^[2]^. The goal of RP is complete prostate extirpation, and despite the favorable cancer control that has been associated with RP, approximately 20-40% of patients experience biochemical recurrence (BCR) ^[3]^. A number of factors have been found to be associated with BCR after RP, and one important adverse risk factor is the presence of a positive surgical margin (PSM) ^[4]^.

Despite the improvements in surgical techniques, equipment, and staging of newly diagnosed patients with PCa that have been attained since the introduction of serum prostate-specific antigen (PSA) screening, 11-38% of patients after RP still have PSM ^[5,6]^. Overall, nearly 30% of the patients with PSM develop BCR, as opposed to approximately 10% of those with negative surgical margins ^[7–9]^.

Considering the significant effect of PSM on postoperative BCR in patients with PCa, the postoperative margin status is an important reference index for adjuvant treatment decisions after RP. The European Association of Urology (EAU) and the National Comprehensive Cancer Network (NCCN) both recommend adjuvant radiotherapy when PSM is present ^[10,11]^. The primary Gleason grade (PGG) prior to PSM, the length of PSM, number of PSMs, and pathological stage of the primary tumor are very important features of PCa with PSM, but their predictive value for BCR in the PSM cohort remains controversial. Therefore, to further clarify the effects of these features, we conducted a meta-analysis of all published epidemiological studies.

## Materials and methods

### Protocol and registration

The results of the systematic review and meta-analysis followed the Preferred Reporting Items for Systematic Reviews and Meta-Analyses (PRISMA) 2020 statement. The study protocol was registered in PROSPERO database (CRD42021255447). See **Texts S1 and S2** for the PRISMA Checklist.

### Search strategy

This meta-analysis was conducted in accordance with the Preferred Reporting Items for Systematic Reviews and Meta-analyses (PRISMA) guidelines. A systematic literature search was performed using electronic databases, including PubMed, EMBASE, and Cochrane Library,from January 1, 2005, to October 1, 2023. The following terms were used for the literature search: (“prostate cancer” OR “prostatic neoplasms”) AND (“prostatectomy”) AND (“positive margin” OR “positive surgical margin” OR “surgical margin”). The search strategies used for English databases are shown in the **S3 Text**.

### Inclusion and exclusion criteria

Eligible studies were determined using the Population, Intervention, Comparator, Outcome and Study design (PICOS) approach. Prospective or retrospective cohort studies were considered eligible if they investigated PGG at PSM, the length of PSM, PSM focality, and pathologic stage of the tumor (C) in patients with PCa with PSMs (P) after RP treatment (I) to assess the independent predictors of BCR (O) using multivariate Cox regression analysis (S).

We excluded reviews, meeting abstracts, case reports, editorials, replies from the authors, letters, and studies without sufficient data. In the case of duplicate publications, either the higher quality or the most recent publication was selected. The references of the included reports were scanned for additional studies. Any disagreements were resolved by discussion with a third investigator.

### Data extraction

Two investigators (HG and YS) independently extracted information from the eligible studies. The following data were extracted: first author’s name, publication year, recruitment country, recruitment period, age, PSA level, number of patients, number of patients with positive surgical margins, follow-up duration, and BCR rate. All discrepancies in data extraction were resolved by discussion between the two reviewers or by consultation with a third reviewer (LZ).

### Quality assessment

The Newcastle-Ottawa Scale (NOS) was used to assess the quality of the enrolled nonrandomized studies. Each study was assessed using eight methodological items, with scores ranging from 0 to 9. Studies with scores of six or higher were graded as high quality. Only high-quality studies were included in further analysis, to ensure the quality of the meta-analysis.

### Statistical analysis

Pooled hazard ratios (HRs) with 95% confidence interval (CIs) were used to evaluate the association between the PGG at PSM, length of PSM, number of PSMs, and pathological stage of the primary tumor. Heterogeneity between studies was evaluated using Cochran’s Q test and the I^2^ statistic. An I^2^ >50% or p-value < 0.1, as determined by Cochran’s Q test, implies that heterogeneity exists. We used a random-effects (RE) model to calculate the pooled HRs for heterogeneous results; otherwise, a fixed-effects (FE) model was adopted. A sensitivity analysis was performed by sequentially excluding each study, to test the reliability of the pooled results. In addition, the presence of publication bias was evaluated using both the funnel plot and Egger’s test. A meta-analysis was performed using the Review Manager software (version 5.3) and STATA 12.0. Statistical significance was set at P < 0.05.

## Results

### Search results

A total of 6,370 potential studies were retrieved and screened according to the retrieval requirements. After duplicates were removed, 4,658 papers remained. After screening, 30 English articles were included in the present meta-analysis, which did not contain any topics, reviews, case reports, republications, or research purposes that were inconsistent with this meta-analysis. Fig. 1 presents the flowchart of the study selection process.

**Fig.1.**
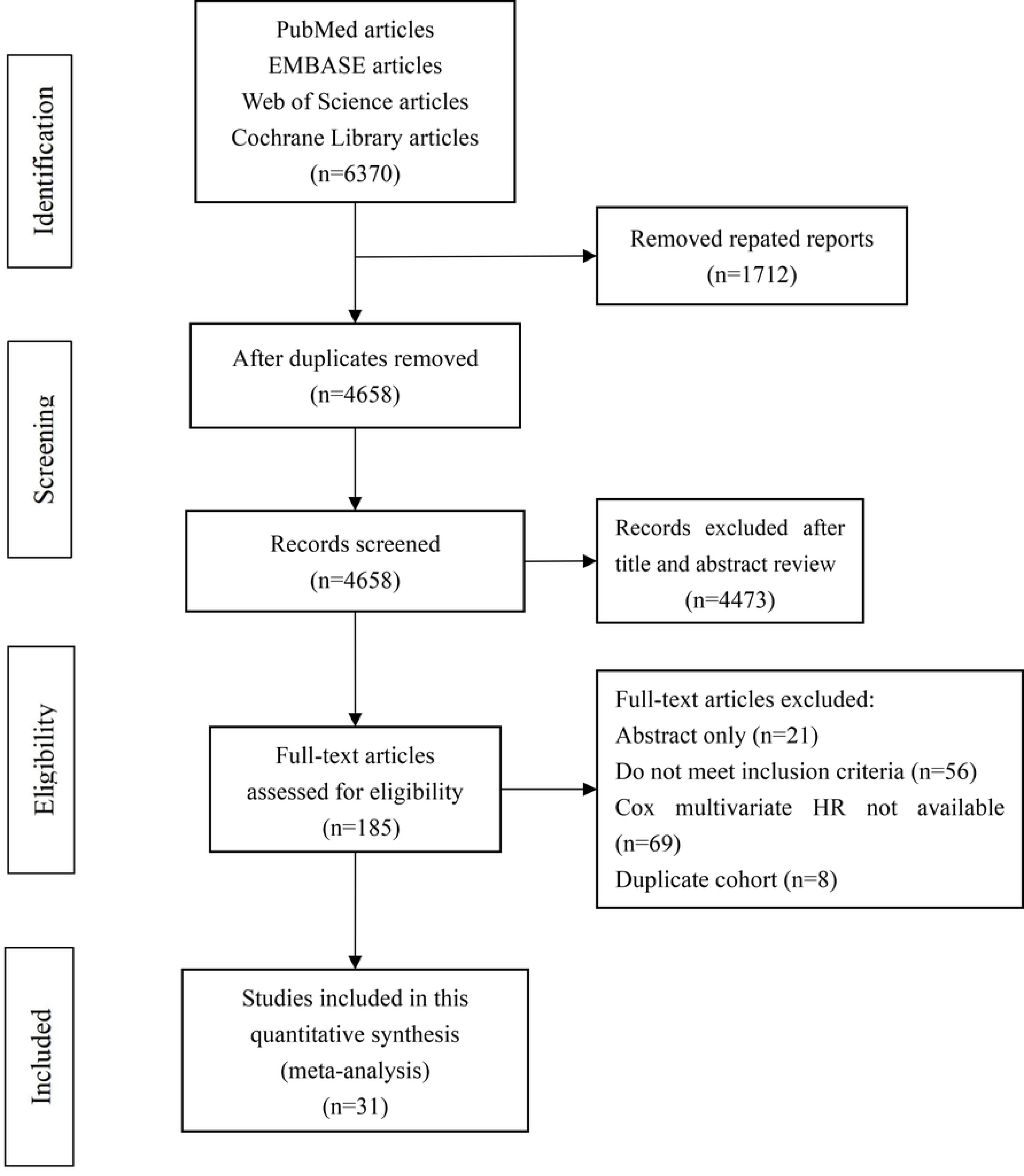
Flowchart of the literature review process for the selection of eligible literatures.

### Study characteristics and quality assessments

The detailed characteristics of the included studies are listed in Table 1.

**Table 1.**
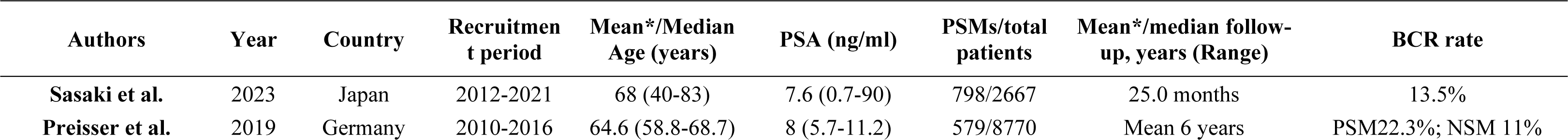
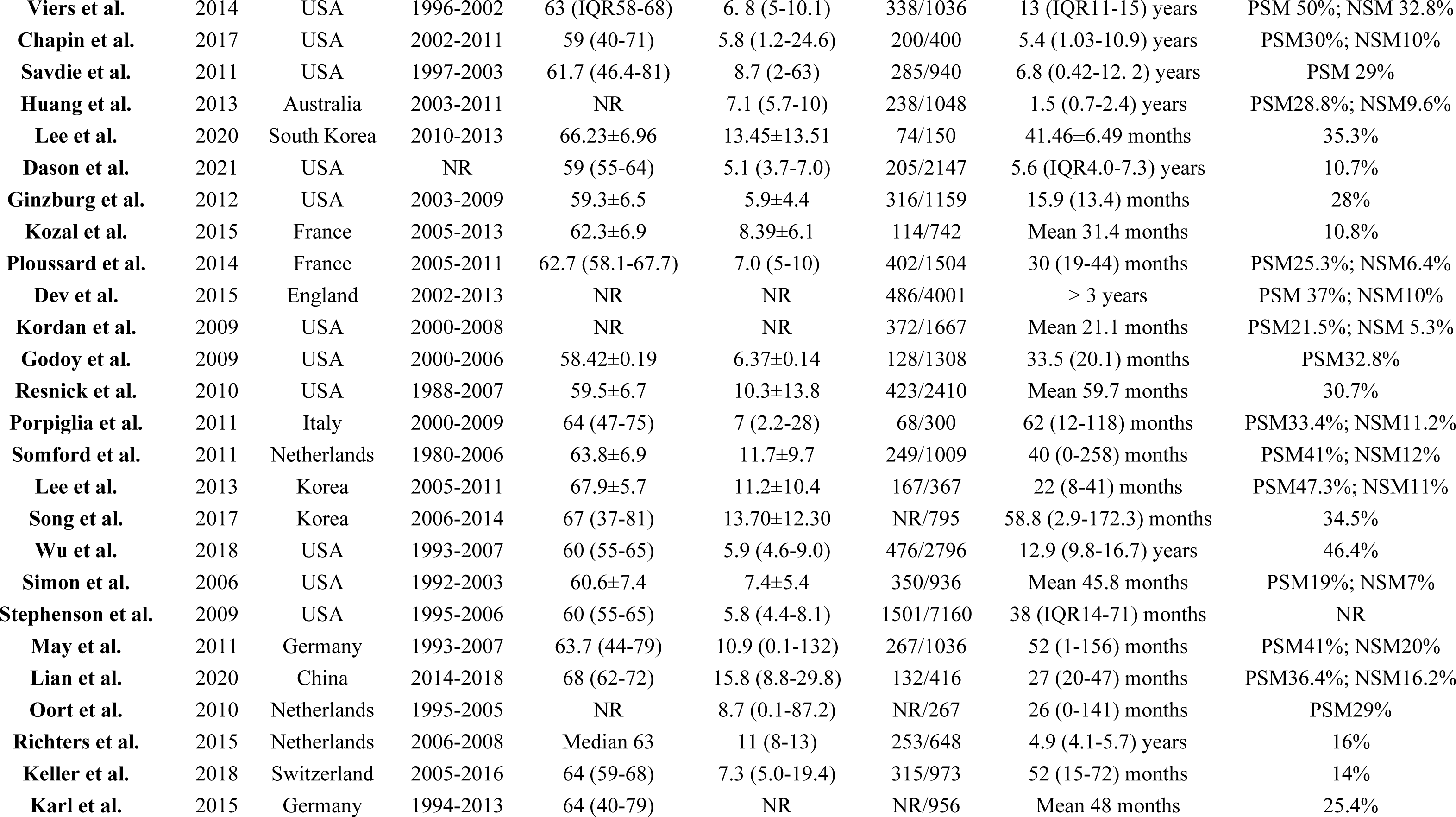
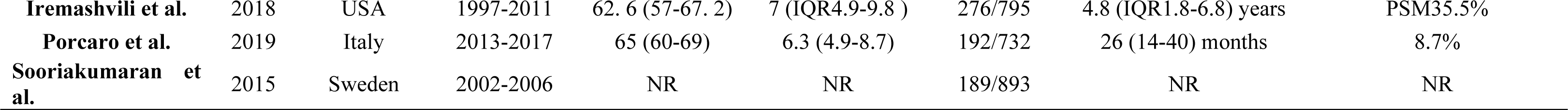
Characteristics of the included studies.

| Authors | Year | Country | Recruitmen<br>t period | Mean*/Median<br>Age (years) | PSA (ng/ml) | PSMs/total<br>patients | Mean*/median follow-up, years (Range) | BCR rate |
| --- | --- | --- | --- | --- | --- | --- | --- | --- |
| Sasaki et al. | 2023 | Japan | 2012-2021 | 68 (40-83) | 7.6 (0.7-90) | 798/2667 | 25.0 months | 13.5% |
| Preisser et al. | 2019 | Germany | 2010-2016 | 64.6 (58.8-68.7) | 8 (5.7-11.2) | 579/8770 | Mean 6 years | PSM22.3%; NSM 11% |
| **Viers et al.** | 2014 | USA | 1996-2002 | 63 (IQR58-68) | 6.8 (5-10.1) | 338/1036 | 13 (IQR11-15) years | PSM 50%; NSM 32.8% |
| **Chapin et al.** | 2017 | USA | 2002-2011 | 59 (40-71) | 5.8 (1.2-24.6) | 200/400 | 5.4 (1.03-10.9) years | PSM30%; NSM10% |
| **Savdie et al.** | 2011 | USA | 1997-2003 | 61.7 (46.4-81) | 8.7 (2-63) | 285/940 | 6.8 (0.42-12.2) years | PSM 29% |
| **Huang et al.** | 2013 | Australia | 2003-2011 | NR | 7.1 (5.7-10) | 238/1048 | 1.5 (0.7-2.4) years | PSM28.8%; NSM9.6% |
| **Lee et al.** | 2020 | South Korea | 2010-2013 | 66.23±6.96 | 13.45±13.51 | 74/150 | 41.46±6.49 months | 35.3% |
| **Dason et al.** | 2021 | USA | NR | 59 (55-64) | 5.1 (3.7-7.0) | 205/2147 | 5.6 (IQR4.0-7.3) years | 10.7% |
| **Ginzburg et al.** | 2012 | USA | 2003-2009 | 59.3±6.5 | 5.9±4.4 | 316/1159 | 15.9 (13.4) months | 28% |
| **Kozal et al.** | 2015 | France | 2005-2013 | 62.3±6.9 | 8.39±6.1 | 114/742 | Mean 31.4 months | 10.8% |
| **Ploussard et al.** | 2014 | France | 2005-2011 | 62.7 (58.1-67.7) | 7.0 (5-10) | 402/1504 | 30 (19-44) months | PSM25.3%; NSM6.4% |
| **Dev et al.** | 2015 | England | 2002-2013 | NR | NR | 486/4001 | > 3 years | PSM 37%; NSM10% |
| **Kordan et al.** | 2009 | USA | 2000-2008 | NR | NR | 372/1667 | Mean 21.1 months | PSM21.5%; NSM 5.3% |
| **Godoy et al.** | 2009 | USA | 2000-2006 | 58.42±0.19 | 6.37±0.14 | 128/1308 | 33.5 (20.1) months | PSM32.8% |
| **Resnick et al.** | 2010 | USA | 1988-2007 | 59.5±6.7 | 10.3±13.8 | 423/2410 | Mean 59.7 months | 30.7% |
| **Porpiglia et al.** | 2011 | Italy | 2000-2009 | 64 (47-75) | 7 (2.2-28) | 68/300 | 62 (12-118) months | PSM33.4%; NSM11.2% |
| **Somford et al.** | 2011 | Netherlands | 1980-2006 | 63.8±6.9 | 11.7±9.7 | 249/1009 | 40 (0-258) months | PSM41%; NSM12% |
| **Lee et al.** | 2013 | Korea | 2005-2011 | 67.9±5.7 | 11.2±10.4 | 167/367 | 22 (8-41) months | PSM47.3%; NSM11% |
| **Song et al.** | 2017 | Korea | 2006-2014 | 67 (37-81) | 13.70±12.30 | NR/795 | 58.8 (2.9-172.3) months | 34.5% |
| **Wu et al.** | 2018 | USA | 1993-2007 | 60 (55-65) | 5.9 (4.6-9.0) | 476/2796 | 12.9 (9.8-16.7) years | 46.4% |
| **Simon et al.** | 2006 | USA | 1992-2003 | 60.6±7.4 | 7.4±5.4 | 350/936 | Mean 45.8 months | PSM19%; NSM7% |
| **Stephenson et al.** | 2009 | USA | 1995-2006 | 60 (55-65) | 5.8 (4.4-8.1) | 1501/7160 | 38 (IQR14-71) months | NR |
| **May et al.** | 2011 | Germany | 1993-2007 | 63.7 (44-79) | 10.9 (0.1-132) | 267/1036 | 52 (1-156) months | PSM41%; NSM20% |
| **Lian et al.** | 2020 | China | 2014-2018 | 68 (62-72) | 15.8 (8.8-29.8) | 132/416 | 27 (20-47) months | PSM36.4%; NSM16.2% |
| **Oort et al.** | 2010 | Netherlands | 1995-2005 | NR | 8.7 (0.1-87.2) | NR/267 | 26 (0-141) months | PSM29% |
| **Richters et al.** | 2015 | Netherlands | 2006-2008 | Median 63 | 11 (8-13) | 253/648 | 4.9 (4.1-5.7) years | 16% |
| **Keller et al.** | 2018 | Switzerland | 2005-2016 | 64 (59-68) | 7.3 (5.0-19.4) | 315/973 | 52 (15-72) months | 14% |
| **Karl et al.** | 2015 | Germany | 1994-2013 | 64 (40-79) | NR | NR/956 | Mean 48 months | 25.4% |
| Iremashvili et al. | 2018 | USA | 1997-2011 | 62.6 (57-67.2) | 7 (IQR4.9-9.8) | 276/795 | 4.8 (IQR1.8-6.8) years | PSM35.5% |
| Porcaro et al. | 2019 | Italy | 2013-2017 | 65 (60-69) | 6.3 (4.9-8.7) | 192/732 | 26 (14-40) months | 8.7% |
| Sooriakumaran et al. | 2015 | Sweden | 2002-2006 | NR | NR | 189/893 | NR | NR |

A total of 46,572 patients (range, 150–8770) with localized PCa after radical prostatectomy were included, in which the PSM samples ranged from 68 to 1,501 (of which 9,836 patients were reported to have PSMs). All 30 studies were retrospective in design, the time of publication ranged from 2006 to 2023, and the mean follow-up durations varied from 1.5 to 13 years. Among the studies, nine^[9,12-19]^, six ^[17,20-24]^, 14 ^[12,17,20,23-33]^ and, 13 ^[23,26-28,30,33-40]^ studies evaluated the relationships between PGG, PSM length, number of PSMs, pathologic stage of the tumor, and BCR, respectively.

NOS scores ranged from 6 to 9, indicating a moderate to high quality of the included studies (**S1 Table**).

### Meta-analysis

Our meta-analysis included all studies that analyzed the relationship between PGG at PSM, the length of PSM, the number of PSMs, pathologic stage of the tumor, and BCR in patients with PCa. Subgroup analyses were conducted according to PGG at PSM.

### Primary Gleason grade of the positive surgical margin

Nine studies including 16,242 patients with PCa compared their BCR between PGG3 and PGG4/5, with HRs with 95% CIs from the multivariate analysis (Table 1). The pooled results, based on the FE model, indicated that compared to PGG3, PGG4/5 was associated with a significantly increased risk of BCR (pooled HR 1.61; 95% CI: 1.34-1.93, P <0.001, I^2^ = 45%; Fig. 2A). A sensitivity analysis was performed by excluding one study at a time, and the results showed that the combined HRs ranged from 1.51 (95% CI: 1.24-1.83) to 1.73 (95% CI: 1.39-2.16) (S1 Fig). Furthermore, the sensitivity analysis did not find any studies that significantly affected heterogeneity (Additional file 1), and the funnel plot did not identify any specific study over the pseudo 95% CI (Fig. 3A). These results indicate that the findings are reliable.

**Fig. 2.**
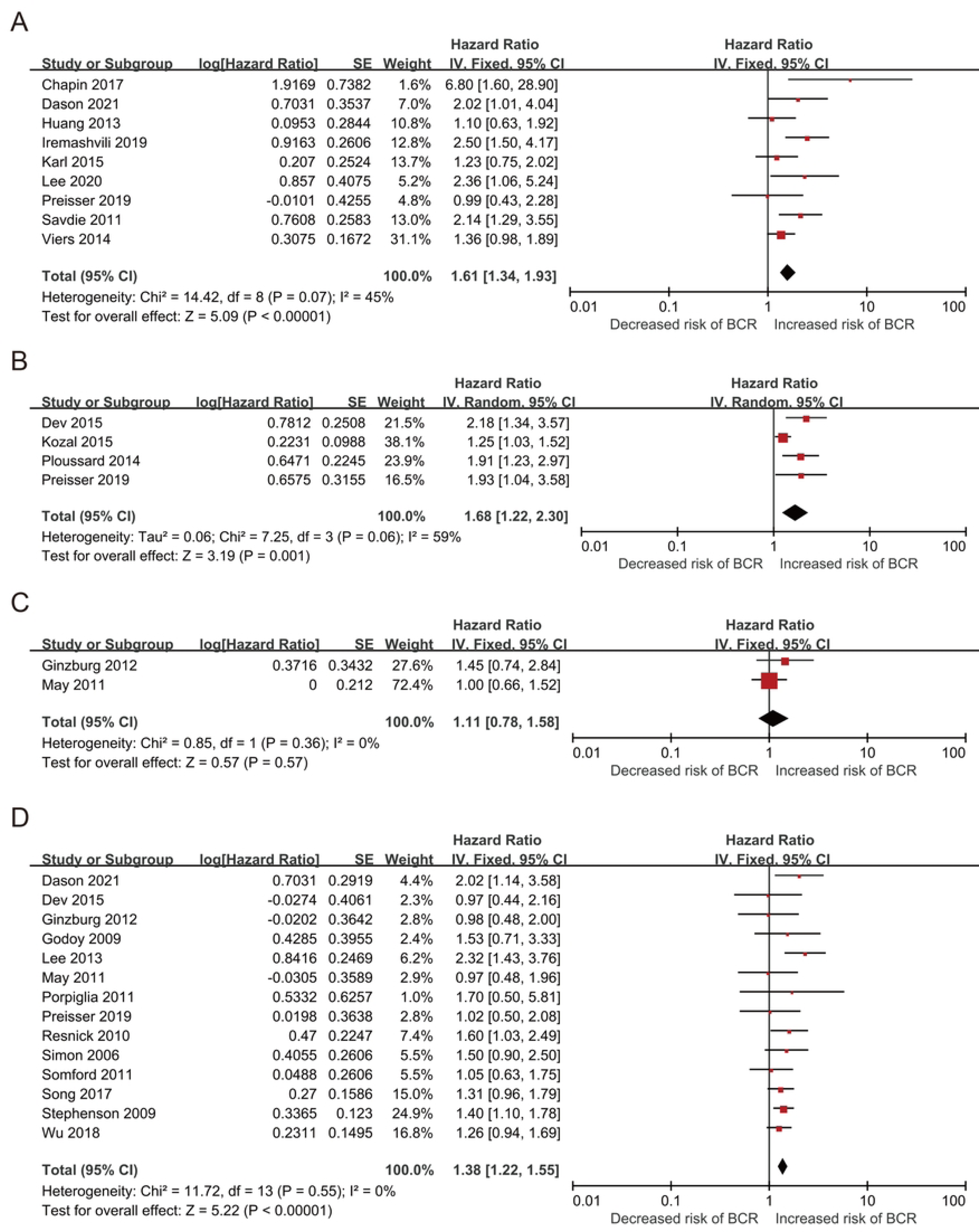
Forest plots of included studies evaluating the association between (A) PGG and BCR, (B)&(C) length of PSM (D) number of PSM in PCa patients.

**Fig. 3.**
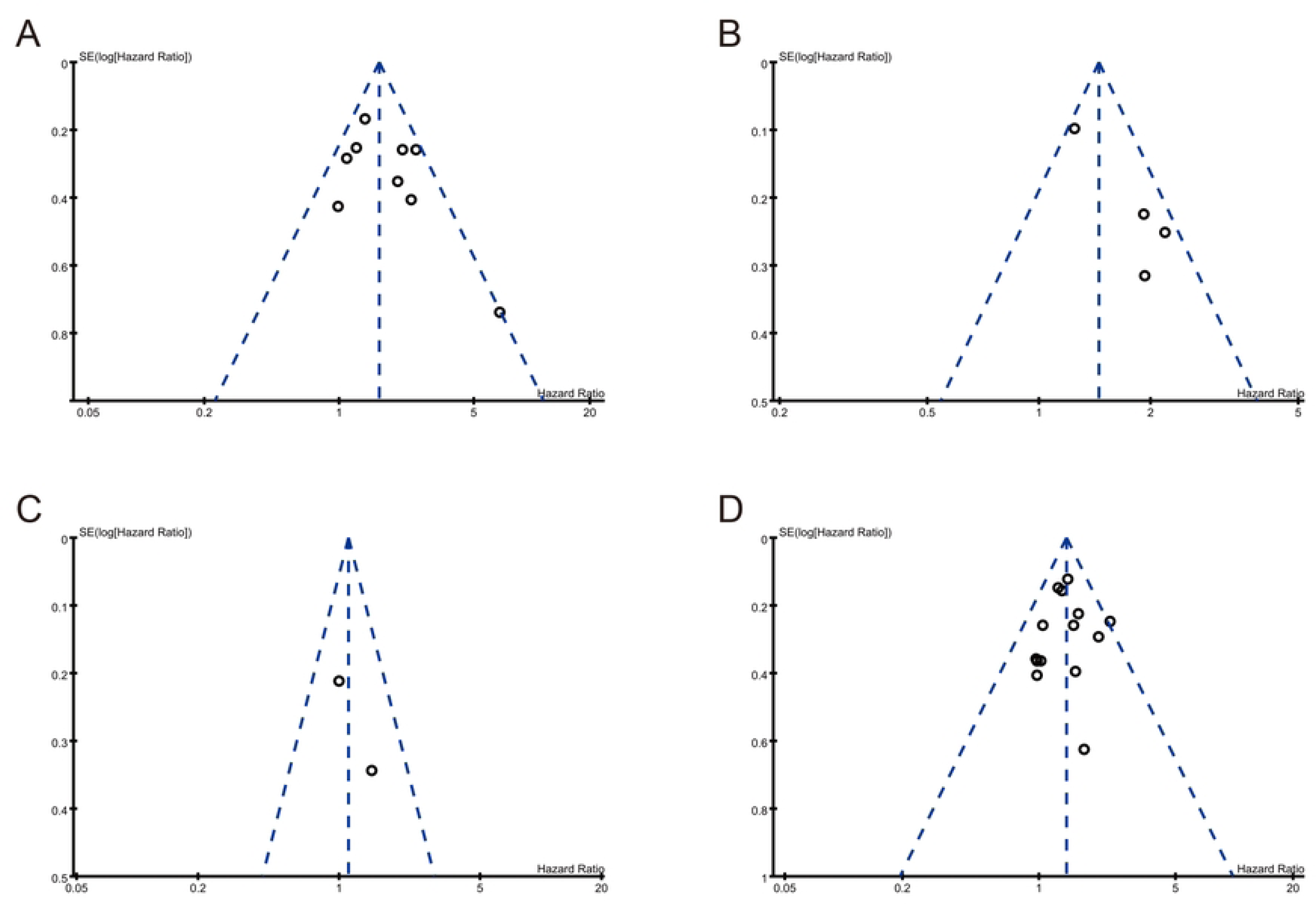
Funnel plots of (A) PGG and BCR, (B)&(C) length of PSM (D) number of PSM and BCR.

### Length of positive surgical margin

Six studies that included 8,592 patients with PCa compared BCR based on the length of PSM <3 mm and PSM ≥3 mm or PSM ≤3 mm and PSM >3 mm, with HR and 95% CIs determined from multivariate analysis (Table 1). Four studies that included 6,397 patients with PCa were classified according to PSM <3 mm and PSM ≥3 mm. The pooled results, based on the random effects model, indicated that compared to PSM < 3 mm, PSM ≥3 mm was associated with a significantly increased risk of BCR (pooled HR 1.68; 95% CI, 1.22-2.30, P=0.001, I^2^=59%; Fig. 2B). Furthermore, the sensitivity analysis showed a significant decrease in heterogeneity (I^2^=0%, P <0.001), after excluding the study by Kozal et al. (Additional File 2). Moreover, the funnel plot identified this study over the pseudo 95% CI (Fig. 3B). After excluding this study, the recalculation results showed a significant correlation between PSM length and BCR without heterogeneity (pooled HR: 2.01, 95% CI: 1.5–2.63; P <0.001; I^2^ = 0%; S2 Fig).

Because the classification methods differ, two articles including 2,195 patients with PCa were classified according to PSM ≤3 mm or >3 mm. The pooled results, based on the FE model, indicated that compared to PSM <3 mm, PSM ≥3 mm was associated with a significantly increased risk of BCR (pooled HR: 1.11; 95% CI: 0.78-1.58, P =0.57, I^2^ = 0%; Fig. 2C).

### Focality of positive surgical margin

Next, we evaluated the relationship between multifocal PSM (mF-PSM) and unifocal PSM, with HR and 95% CIs determined from a multivariate analysis. Fourteen studies with 34,194 patients were evaluated (Table 1). The pooled results, based on the FE model, indicated that compared to unifocal PSM, mF-PSM was associated with a significantly increased risk of BCR (pooled HR 1.38; 95% CI: 1.22-1.55, P <0.001, I^2^ = 0%; Fig. 2D). A sensitivity analysis was performed by excluding one study at a time, and the results showed that the combined HRs ranged from 1.33 (95% CI: 1.18-1.51) to 1.40 (95% CI: 1.23-1.6) (Additional file 3). In addition, the sensitivity analysis did not find a single study that significantly affected heterogeneity (Additional file 3), and the funnel plot did not identify any study with a pseudo 95% CI (Fig. 3D). These results indicate that our findings are robust.

### Pathologic stages of the PSM cohort

Thirteen studies that included 13,637 patients with PCa compared BCR values between different pathological stages, with HRs and their 95% CIs from a multivariate analysis (Table 1). Five studies included 4,502 patients and compared their BCR between pT3 and pT2. The pooled results, based on the random effects model, indicated that compared to pT2, pT3 was associated with a significantly increased risk of BCR (pooled HR: 1.66; 95% CI: 1.06-2.6, P<0.001, I^2^ = 67%; Fig. 4A). Moreover, the funnel plot identified two articles with a pseudo 95% CI (Fig. 5A). Due to the high heterogeneity, we further conducted a subgroup analysis.

**Fig. 4.**
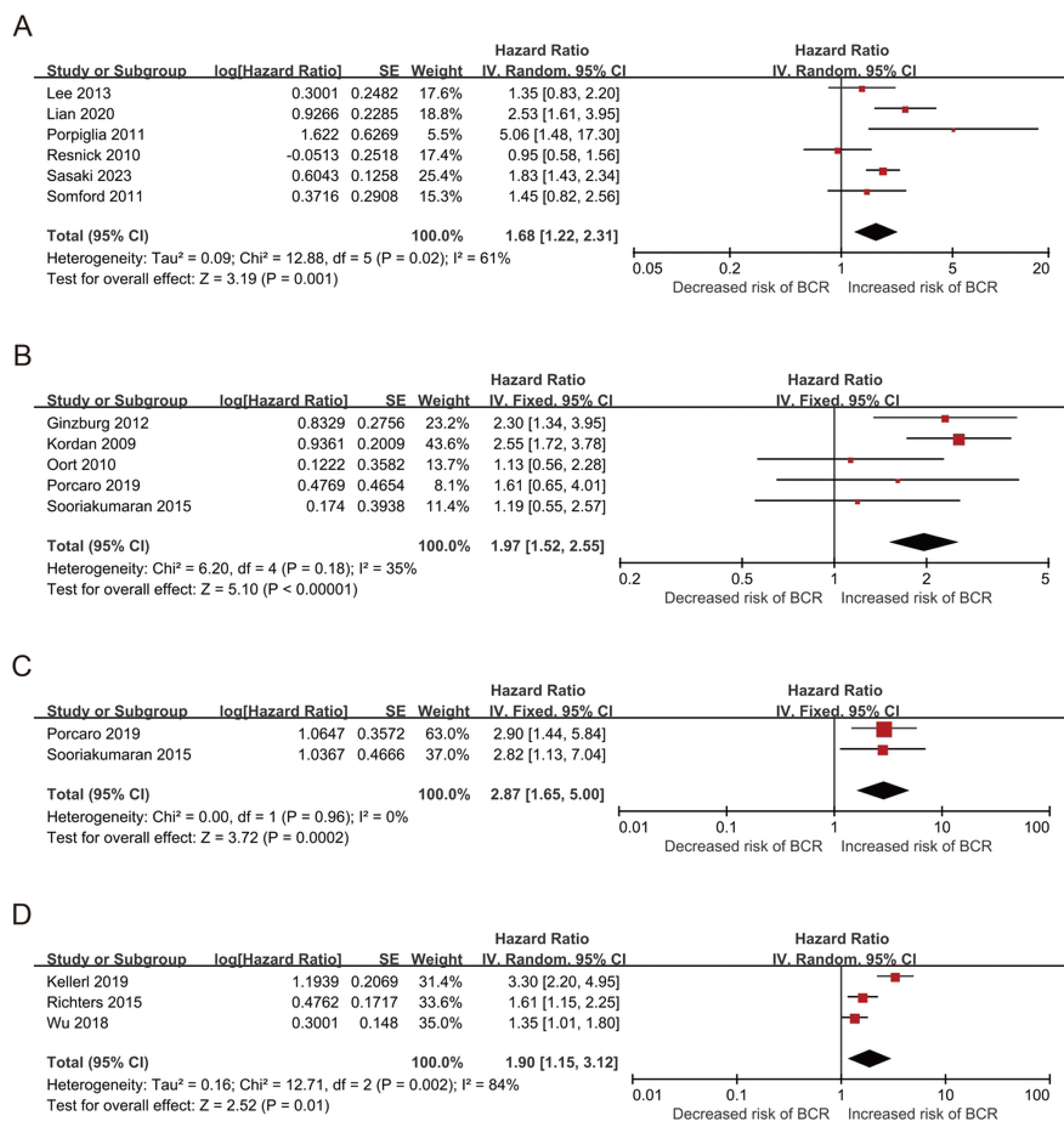
Forest plots of included studies evaluating the association between (A) pT2 and pT3, (B) pT2 and pT3a, (C) pT2 and pT3b, (D) pT2 and pT3/4.

**Fig. 5.**
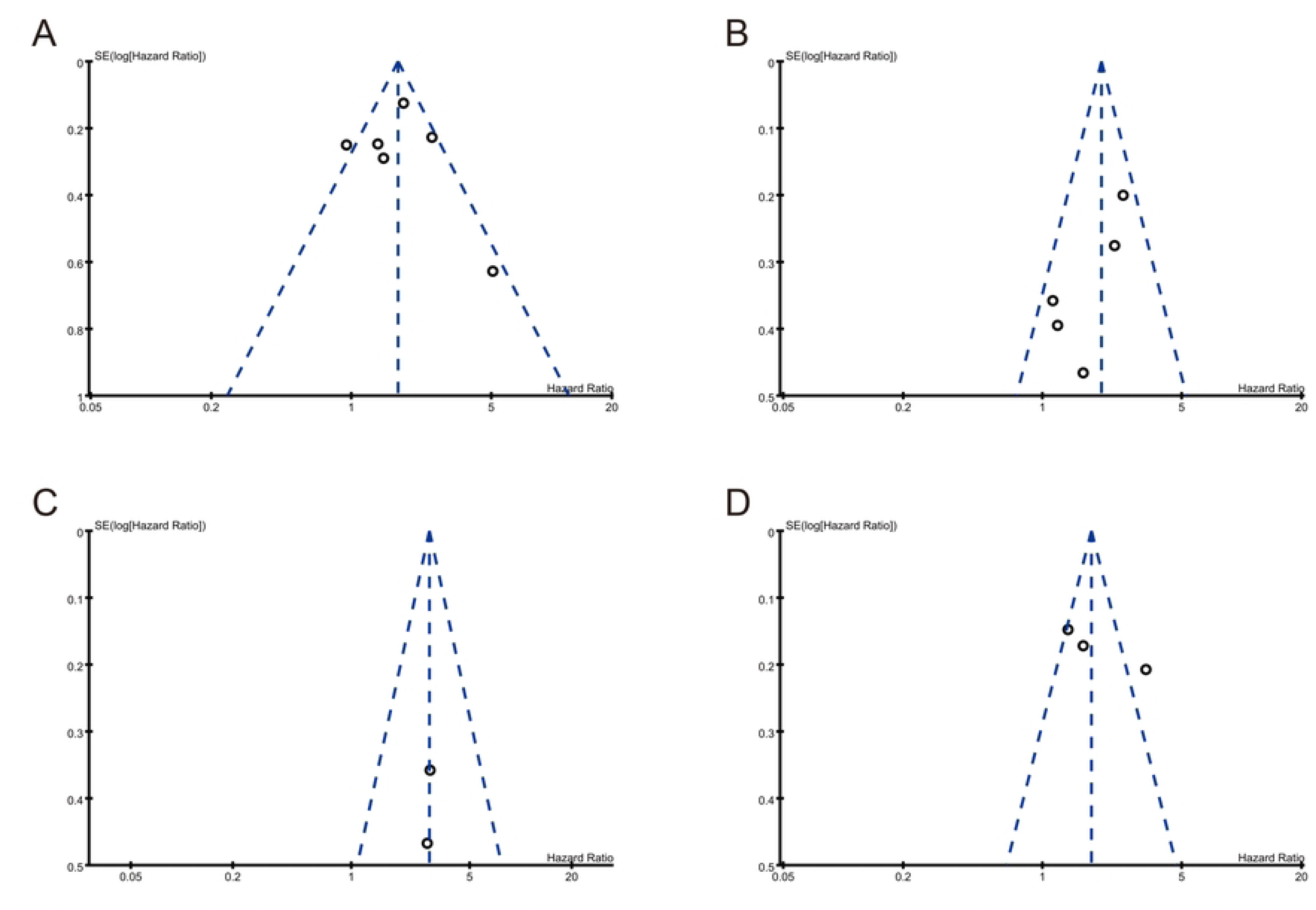
Funnel plots of included studies evaluating the association between pathologic stage of the PSM (A) pT3 and pT2, (B) pT3a and pT2, (C) pT3b and pT2, (D) pT3/4 and pT2.

Next, we evaluated the relationship between pT3a and pT2 using HRs and 95% CIs and included five studies with 4,718 patients. The results showed that based on the FE model, the pooled results indicated that compared to pT2, pT3a was associated with a significantly increased risk of BCR (pooled HR: 1.97; 95% CI: 1.52-2.55; P<0.001; I^2^=35%; Fig. 4B). A sensitivity analysis was also conducted, in which one study was removed at a time, and the results showed that the combined HRs ranged from 1.61 (95% CI: 1.14-2.28) to 2.15 (95% CI: 1.62-2.84) (Additional file 4). Notably, the funnel plot analysis did not identify any study with a pseudo 95% CI (Fig. 5B), which indicates that our findings are reliable.

Furthermore, two studies that included 1,625 patients assessed the association between pT3b and pT2. The synthesized analysis showed that, compared to pT2, pT3b was associated with a significantly increased risk of BCR (pooled HR: 2.87; 95% CI: 1.65-5.00, P <0.001, I^2^ =0%; Fig. 4C). Furthermore, no studies with a pseudo 95% CI were found in the funnel plot analysis (Fig. 5C).

We evaluated the relationship between pT3/4 and pT2 using HR and 95% CIs and included three studies with 4,417 patients. The random-effects model results showed that compared to pT2, pT3/4 was associated with a significantly increased risk of BCR (pooled HR: 1.90; 95% CI: 1.15-3.12, P <0.001, I^2^=84%; Fig. 4D). The sensitivity analysis showed that heterogeneity was evidently reduced (I^2^ = 0%, P <0.001; Additional file 5) after excluding the study by Kellerl et al. The pooled HR, recalculated using the FE model, was 1.46 (95% CI: 1.17-1.81, P <0.001; Additional file 5). Moreover, the funnel plot identified it over the pseudo 95% CI (Fig. 5D).

### Subgroup analysis

Our results confirmed that PGG at PSM was associated with a significantly increased risk of BCR, based on nine studies. However, the heterogeneity remained significant after the sensitivity analysis. As different study features were involved, we further performed a subgroup analysis to explore the source of this heterogeneity (Table 2). Collectively, the results of the subgroup analyses indicated that the median age, region, sample size, and median follow-up months did not affect the relationship between PGG and BCR in patients with PCa, whereas the sample size and median age could be a potential source of heterogeneity.

**Table 2.** Subgroup analysis for primary Gleason grade (PGG) at PSM.

| Subgroup | No. of studies | No. of patients | HR (95% CI) | P value | Heterogeneity |  | Effect model |
| --- | --- | --- | --- | --- | --- | --- | --- |
| | | | | | $I^2$ (%) | Ph | |
| **Overall** | 9 | 16242 | 1.61 (1.34-1.93) | <0.001 | 45% | 0.07 | fixed |
| **Region** |  |  |  |  |  |  |  |
| Europe | 8 | 16092 | 1.57 (1.3-1.9) | <0.001 | 48% | 0.06 | fixed |
| Asia | 1 | 150 | - | - | - | - | - |
| **Sample size** |  |  |  |  |  |  |  |
| <1000 | 5 | 3241 | 1.93 (1.45-2.58) | <0.001 | 43% | 0.13 | fixed |
| ≥1000 | 4 | 13001 | 1.33 (1.04-1.71) | 0.02 | 0% | 0.5 | fixed |
| **Median age (years)** |  |  |  |  |  |  |  |
| ≥63 | 4 | 10912 | 1.36 (1.06-1.74) | 0.01 | 0% | 0.47 | fixed |
| <63 | 5 | 4282 | 2.36 (1.73-3.23) | <0.001 | 0% | 0.49 | fixed |
| **Median time of follow-up (years)** |  |  |  |  |  |  |  |
| <4.8 | 3 | 2154 | 1.32 (0.95-1.85) | 0.1 | 20% | 0.29 | fixed |
| ≥4.8 | 6 | 14088 | 1.74 (1.4-2.17) | <0.001 | 50% | 0.07 | fixed |

### Publication Bias

According to the funnel plot analysis, most of the studies were located at the upper part of the “inverted funnel” with a roughly symmetrical distribution, which indicated that the included studies had no significant publication bias. Publication bias was also assessed using Egger’s test. The results showed no significant publication bias in PGG (P = 0.215), length of PSM (P=0.052), PSM focality (P=0.815), and pathologic stage of the tumor (P=0.155, 0.075, 0.556, respectively), indicating the robustness of the results.

## Discussion

Due to the influence of prostate anatomy and other factors, the positive rate of surgical margins after RP ranged as high as 11% to 38%, ranking first among malignant tumors in men ^[5,6]^. Prior studies have shown that PSM is associated with BCR after RP, and the EAU and NCCN recommend adjuvant radiotherapy for RP patients with PSM after RP ^[10,11]^. However, because the side effects of radiotherapy and the effect of radiotherapy on patient long-term survival have not been determined ^[41,42]^, the proportion of patients with PSM who actually received adjuvant radiotherapy after surgery was relatively low ^[43]^.

To identify patients with different risks of BCR and to more accurately select patients who have the greatest opportunity to benefit from postoperative therapy, it is necessary to perform risk stratification in the study patients with PSM. Notably, this may limit potential overtreatment and reduce medical costs. Due to differences in genetic and environmental background, detection means, operation methods, and surgical techniques, the prognosis of PCa heterogeneity cannot be fully elucidated. Only multicenter studies that include a large number of events involving different genetic or environmental backgrounds can reasonably evaluate the factors in predicting clinical outcomes in the PSM cohort. Thus, the aim of our systematic review and meta-analysis of 30 studies involving 46,572 patients with PSM (21.1%) was to summarize and analyze the current evidence regarding the predictive value of the PSM parameters and primary tumor pathological stage on BCR in patients with PCa. The results showed that compared to PGG3, PSM < 3 mm, single PSM focality, lower pathologic stage, PGG ≥ 3, PSM ≥ 3 mm, multiple PSM focality, and higher pathologic stage were associated with a significantly increased risk of BCR. The sensitivity and subgroup analyses further revealed the reliability and rationality of our findings. Collectively, the pooled data from this meta-analysis confirmed that the PGG at the PSM, its length, PSM focality, and pathologic stage of the tumor could predict clinical outcomes and may serve as reliable prognostic indicators for patients with PCa complicated with PSM after RP treatment.

The Gleason score of the primary tumor is known to be an extremely important pathological parameter of PCa and was found to be significantly associated with BCR ^[13,18,19,44,45]^. However, the predictive value of the Gleason score in PSM remains controversial ^[9,13,16-18,45]^. Hollemans et al. ^[45]^ found that up to 54% of the Gleason grade of tumors at the PSM differed from that of primary tumors. Notably, the Gleason grade of tumors at the PSM was found to be even better than the Gleason grade of the primary tumor in predicting BCR in the multivariate analysis. We performed a meta-analysis of nine studies involving 16,242 patients with PCa, and the pooled results showed that PGG4 or 5 was associated with a significantly increased risk of BCR compared to PGG3. However, after the sensitivity analysis, heterogeneity remained significant. Because different study characteristics were involved, we performed a subgroup analysis to explore the source of this heterogeneity. The results showed that sample size and median age could be potential sources of heterogeneity. In conclusion, an elevated Gleason grade on PSM was associated with an increased risk of BCR.

Interestingly, the effect of Gleason grade on PSM was independent of the predominant Gleason pattern in the tumor, suggesting a differential biological effect. The underlying mechanism could be that the wound environment after prostatectomy leads to the production of many cytokines and angiogenic factors that respond differently to residual cancers of different Gleason grades ^[13]^.

Possible factors include vascular endothelial growth factor (VEGF) and VEGF receptor fetal liver kinase (Flk)-1, which are expressed at high levels in wounds. Both can not only induce the proliferation of vascular endothelial cells but also directly induce the proliferation of PCa cells, which increase in PCa in a grade-dependent manner. Similar findings have been reported for basic fibroblast growth factor (bFGF), fms-like tyrosine kinase 1 (Flt-1), TGF-β receptors, fibroblast growth factor, and platelet-derived growth factor ^[13,46,47]^.

Contrary to the results of other studies, Huang et al. ^[13]^ found that the presence of PGG4 / 5 at the pathological margin had no significant association with the risk of BCR. However, when the PSADT was ≤ 9 months and ≤ 6 months, the presence of Gleason pattern 4 / 5 at the pathological margin was an independent predictor of BCR. Meanwhile, the follow-up time in this study was relatively short. Notably, in a large study with a median follow-up time of 13 years, Viers et al. ^[19]^ reported that although PGG4 was not significantly associated with the risk of BCR-FS in a multivariable analysis, it was associated with a significantly increased risk of PCa-specific mortality and systemic progression.

Since PSM after RP has been shown to be associated with an increased incidence of BCR and mF-PSM should have more residual tumor tissues in the surgical bed, it is speculated that having more than one PSM should reflect a more aggressive disease, leading to increased BCR. Fourteen studies involving 34,194 patients were evaluated, and the pooled results based on a FE model showed that mF-PSM was associated with a significantly increased risk of BCR compared with single-focal PSM, which is consistent with the majority of studies included.

In contrast, Song ^[31]^ and Wu ^[33]^ retrospectively analyzed 821 and 476 patients with PCa with PSM after RP and found that patients with mF-PSM showed significantly worse BCR prognosis on univariate analysis. This finding lost significance in the multivariate analysis, in agreement with the findings of several other studies ^[23-25,27,29,30,36]^. There are several possible reasons for this finding. Firstly, mF-PSM is more likely to occur in patients with higher pathological stages and to be associated with higher PSA levels, pT stage, and GS. This suggests that the prognostic impact of mF-PSM is influenced by other high-risk factors. At the same time, the number of patients with mF-PSM was relatively small, and they were more likely to receive adjuvant therapy after surgery, leading to their exclusion from the final study. Furthermore, the association of mF-PSMs with BCR may be influenced by other risk factors. Lastly, in non-organ-confined disease, PSM may occur at a location far from the extraprostatic extension site.

The impact of margin extent is similar to the impact of multifocality on BCR. Concerning the impact of PSM size on BCR, the comparability of various reports is limited by the different thresholds used to differentiate small and large PSM sizes. For reasons of statistical efficiency, studies generally reduce the margin extent into a categorical variable, often separated into <1/≥1 mm or <3/≥3 mm. The results of our meta-analysis of four studies ^[17,20-22]^ showed that PSM ≥ 3 mm was associated with a significantly increased risk of BCR compared with PSM < 3 mm. The sensitivity analysis showed a significant reduction in heterogeneity after the exclusion of the study by Kozal et al. ^[21]^. This may be due to the fact that Kozal’s study selected subjects for prostate resection using a single robot-assisted radical prostatectomy (RARP) method with a short follow-up period. Two studies with 2,195 patients with PCa were classified according to PSM ≤ 3 mm and PSM > 3 mm. The pooled results indicated that compared to PSM < 3 mm, PSM ≥ 3 mm was associated with a significantly increased risk of BCR. This finding indicates that patients with broad-spread PSM will develop a more aggressive disease, leading to decreased BCR-free survival. The primary findings suggest that patients with a PSM length of > 3 mm may be eligible for more aggressive adjuvant therapy. In addition, other classified methods exist, such as PSM > 1 mm vs. PSM ≤ 1 mm, PSM < 2 mm vs. PSM ≤ 2 mm, PSM > 2.8 mm vs PSM ≤ 2.8 mm, and others. Owing to the limited number of studies, no relevant meta-analysis was conducted.

Dev (2015) et al. ^[20]^ reported that both multifocality and PSM ≥3 mm increased the risk of BCR compared with NSM, while in the analysis of the PSM group, only PSM ≥3 mm remained a significant predictor of BCR. Multifocality appears to confer less additional harm in terms of BCR risk, whereas margin length ≥3 mm remains highly predictive of BCR compared to margins < 3 mm. One possible explanation for this phenomenon is that the presence of multiple smaller PSMs (< 3 mm) during RP may reflect iatrogenic lesions, especially if the error has not been detected, rather than a single invasive lesion that breaches the surgical margin.

In contrast to these studies, May et al. ^[24]^ found that although PSM was an independent predictor of BCR, in a further subgroup analysis, the BCR of patients with PSM > 3 mm vs ≤ 3.0 mm did not differ. KARL et al. ^[15]^ conducted a study that included 608 patients with pT3aN0/Nx PCa with PSM. The results showed that, although PSM > 3 mm was an independent predictor of BCR in the univariate analysis, it was not an independent predictor of BCR in the multivariate analysis. Residual tumors are highly suggestive of cancer treatment failure; however, residual tumors of small size may fail to continue to grow due to damage through debris formation and reparative processes, as well as changes in the microenvironment of the tumor body and damage of tumor cells around the cutting edge. In addition, smaller-sized PSMs are more likely to be in the last layer of the tumor, after which the tumor ceases to continue spreading into the adjacent tissue. A higher pathological stage of PCa predicts increased tumor aggressiveness, which has been widely reported to be associated with an increased BCR. However, its effect on the BCR in the PSM cohort remains controversial.

In contrast to previous reports, Somford et al. ^[30]^ reported no significant difference in the BCR of pathological staging in a multivariate analysis. Moreover, Sooriakumaran ^[38]^ and Servoll et al. ^[48]^ reported that, compared to the pT3 cohort, PSM had a greater impact on the pT2 cohort in predicting BCR. A possible explanation is that the patients with a higher pathological stage were more likely to receive adjuvant therapy after surgery and were therefore excluded from the final study cohort. In addition, patients with pT3 status are more likely to be complicated with PSM ^[23,38,48]^. However, in the multivariate analysis, PSM may have a more significant effect on BCR, which would diminish the effect of pathological staging on BCR.

In summary, this meta-analysis revealed that PGG≥3, PSM ≥3 mm, multiple PSM focality, and a higher pathologic stage of the tumor were associated with an increased risk of BCR in patients with PCa with PSM, which may also be associated with higher biological aggressiveness and should be treated differently in terms of adjunct treatment and monitoring frequency. In view of this, we recommend that the Gleason score at the PSM, its length, and PSM focality be routinely reported as part of the standard pathology document. Large-scale and well-designed prospective studies with longer follow-up periods are needed to validate the efficacy of these risk factors and their effect on the response to adjuvant and salvage therapies in this population, as well as on other oncological outcomes.

Nevertheless, the present study had some limitations that should be acknowledged. First, all the included studies were retrospective, which may have led to a selection bias. Second, in this study, there was no subgroup analysis on the impact of the Gleason score at the PSM, its length, and the PSM focality on BCR. This is because there were few relevant studies and inconsistent relevant classification standards. Third, the literature involved in this study was limited to papers published in English, which may have led to selection bias. Finally, heterogeneity was detected among the enrolled studies in some data analyses, which could have influenced the final results.

## Data Availability

A systematic literature search was performed using electronic databases, including PubMed, EMBASE, Cochrane Library, and Web of Science, from January 1, 2005, to October 1, 2023. The protocol was pre-registered in PROSPERO.

## Supporting information

**S1 Table. NOS Scores of included study.(PDF)**

**S1 Fig.(A) Forest plots of studies excluded Iremashvili’s study evaluating the association between PGG and BCR, (B) Forest plots of studies excluded Vier’s study evaluating the association between PGG and BCR.**

**S2 Fig.Forest plots of studies excluded Kozal’s study evaluating the association between length of PSM and BCR.**

**Additional file 1.Forest plots of studies evaluate the association between PGG and BCR.(PDF)**

**Additional file 2.Forest plots of studies evaluate the association between length of PSM and BCR (<3 mm VS≥3 mm).(PDF)**

**Additional file 3.Forest plots of studies evaluate the association between focality of PSM and BCR.(PDF)**

**Additional file 4.Forest plots of studies evaluate the association between pT3a/pT2 and BCR.(PDF)**

**Additional file 5.Forest plots of studies evaluate the association between pT3-4/pT2 and BCR.(PDF)**

**S1 Text. PRISMA Abstract Checklist.(DOCX)**

**S2 Text. PRISMA 2020 Checklist.(DOCX)**

**S3 Text. Search strategy.(DOCX)**

## Acknowledgments

We would like to thank all the participants for their support during the follow-up study, as well as the researchers who had published high-quality articles for our meta-analysis.

## Author Contributions

Conceptualization: Hong Guo,Lei Zhang,Yuan Shao.

Data curation: Hong Guo,Lei Zhang,Kunyang An.

Formal analysis:Hong Guo,Yuan Shao,Kunyang An.

Investigation:Hong Guo,Yuan Shao,Kunyang An.

Methodology:Hong Guo,Lei Zhang,Kunyang An, Caoyang Hu,Dongwen Wang.

Project administration:Caoyang Hu,Xuezhi Liang,Dongwen Wang.

Resources:Hong Guo,Dongwen Wang.

Software:Hong Guo,Lei Zhang,Yuan Shao,Kunyang An.

Supervision:Caoyang Hu,Xuezhi Liang,Dongwen Wang.

Validation:Hong Guo,Yuan Shao,Xuezhi Liang.

Visualization:Lei Zhang.

Writing – original draft:Hong Guo,Lei Zhang,Caoyang Hu.

Writing – review & editing:Dongwen Wang.

